# *APOL1* Risk Variants Associate with the Prevalence of Stroke in African American Current and Past Smokers

**DOI:** 10.1101/2023.04.28.23289292

**Authors:** Jelena Mustra Rakic, Clive R. Pullinger, Erin L. Van Blarigan, Irina Movsesyan, Eveline Oestreicher Stock, Mary J. Malloy, John P. Kane

**Author notes:** **Corresponding author:** Jelena Mustra Rakic, PhD, Cardiovascular Research Institute, Center for Tobacco Control Research and Education, University of California, San Francisco, 555 Mission Bay Boulevard South, Room 282, San Francisco, CA 94158. ***Author Contributions:*** Authors contributed to the manuscript in the following ways: study concept and design (JMR, CRP, JPK, EVB); recruitment of the participants (JPK, MJM, EOS); acquisition and analysis of the specimens (JMR, CRP, IM); drafting the manuscript (JMR); critical revision of the manuscript for important intellectual content (JMR, CRP, JPK, EVB, EOS, MJM, IM); collecting, statistical analysis and interpretation of the data (JMR, CRP, EVB, JPK, EOS); obtained funding (JMR, JPK). All authors have approved the final article. **Emails:** JMR CRP ELVB IM EOS MJM JPK.

## Abstract

**Introduction:** Among African Americans, tobacco smokers have 2.5 times higher risk for stroke compared to non-smokers; the tobacco-related stroke risk being higher than in other races/ethnicities. About one half of African Americans carry at least one of two genetic variants (G1 and G2; rare in other races) of apolipoprotein L1 (apoL1), a component of high-density lipoproteins. Several studies showed *APOL1* G1/G2 risk variants associate with stroke. However, the role of *APOL1* variants in tobacco-related stroke is unknown.

**Methods:** In a cross-sectional study, we examined whether *APOL1* risk variants modify the relationship between smoking and stroke in 513 African American adults (median age 58 years, 52% female) recruited through the University of California, San Francisco Lipid Clinic. Using DNA, plasma, and questionnaires we determined *APOL1* variants, smoking status, and history of stroke. Using unstratified and stratified multivariable logistic regression models we examined the association between smoking history (*ever* smokers vs. *never* smokers) and odds of stroke overall, and among carriers of risk variants and non-carriers, separately.

**Results:** Among participants, 41% were *ever* (*current* and *past*) smokers, 54% were carriers of the *APOL1* risk variant, and 41 have had stroke. In all stroke cases, where full medical records were available, stroke types were determined to be an ischemic, and not hemorrhagic, stroke. The association of smoking history and stroke differed by *APOL1* genotype status in the unstratified model (*P*^interaction term^=0.016). Among carriers of risk variants, *ever* smokers had odds ratio (OR) =2.88 for stroke compared to *never* smokers (P=0. 0.038). The OR for stroke comparing *ever* vs. *never* smokers showed a dose-response trend among carriers of one risk allele of 2.35 and two risk alleles of 4.96. Among non-carriers, smoking history was not associated with a stroke.

**Conclusion:** In conclusion, current and past smokers who carry *APOL1* G1 and/or G2 risk variants may be more susceptible to stroke, in particular ischemic stroke, among African Americans.

## INTRODUCTION

African American/Black people (African American) have a disproportionately higher risk for stroke compared to all other races in the United States, which is only partially explained by differences in traditional risk factors and sociological inequalities.^1^ Tobacco smoking, a well-known independent risk factor for stroke, has been shown to be responsible for roughly one fifth of all strokes in United States.^2,3^ Yet, limited data are available on the relationship between smoking and incident of stroke among African American people. Recently, in a prospective cohort study, African American current smokers were found to have 2.5 times higher risk for stroke compared to African American non-smokers.^4^ In contrast, in the Framingham Heart Study, investigators have reported risk ratio for stroke to be ∼1.5 in people who identify as non-Hispanic White.^4,5^ The greater risk of stroke due to tobacco smoking among African American people, compared to people who identify with other races,^4^ underscores the necessity to better understand the etiology of this relationship.

Much of what we know about traditional risk factors for cardiovascular diseases, including stroke, such as systolic hypertension, diabetes mellitus, body mass index, tobacco smoking, lipid profile, has come from the Framingham Heart Study, a prospective cohort study that has been conducted with White individuals of European descent in one geographic area of the United States.^5-8^ This limits our ability to identify risk factors beyond those characteristics for White people of European descent.

Two clinically important variants in the *APOL1* gene (G1 and G2) are common in African American people, and extremely rare in White people.^9^ Due to the slave-trade and forced migration of people from the Atlantic coast of Africa to America, the *APOL1* G1 and G2 frequency among African American people in the United States is high, with approximately 50% carrying at least one risk allele.^9-11^ The *APOL1* gene encodes the apoL1 protein, a component of high-density lipoprotein (HDL) particles.^10^ ApoL1 has an important function in the innate immune responses to *Trypanosoma brucei*.^9^ The G1 and G2 variants in the terminal exon of the *APOL1* may offer enhanced protection against *Trypanosoma brucei*.^*12*^ Additionally, the presence of *APOL1* G1 and/G2 variants is associated with increased risk for kidney disease, with odds ratios of 3–29 among populations of sub-Saharan African descent.^9,12^

*APOL1* G1 and G2 variants explain a substantial fraction of the excess risk for non-diabetic kidney disease in African American people.^9^ Interestingly, several studies examining a relationship between these variants and risk of stroke reported an increased risk for stroke among carriers of the *APOL1* G1 and G2 variants.^10,11,13^ However, other studies failed to find an association.^14-16^ In a transgenic mice study, a biologic mechanism involved in kidney disease development in carriers of the *APOL1* risk genotype was found to depend not only on presence of G1 and G2 risk variants, but also on risk variant expression levels,^17^ suggesting an increase of apoL1 levels to be “second hit” needed for disease development. Given that tobacco smoking-related inflammation may increase the levels of apoL1,^18^ which has been shown to accumulate in coronary plaque of *APOL1* G1 and/or G2 variant carriers leading to its rupture,^19^ unraveling the role of these variants among African Americans in tobacco related-stroke is of immense significance for understanding and preventing the greater burden of tobacco-related stroke in this high-risk population. In our cross-sectional study, we assessed whether the presence of *APOL1* genotypes modified the association between tobacco smoking and stroke prevalence among 527 African American adults.

## MATERIALS AND METHODS

### Study participants

A total of 513 self-reported African Americans (52% female) were included in the study. Participants attending the University of California, San Francisco (UCSF) Lipid and Endocrinology Clinics between 1999 and 2019 were recruited into the UCSF Genomic Resource in Arteriosclerosis (GRA) study.^20^ All participants gave written informed consent prior to their enrollment in the study, which adhered to the World Medical Association Declaration of Helsinki. The UCSF Institutional Review Board as part of the UCSF Human Research Protection Program approved the study. Children were not included. The data that support the findings of this study are available from the corresponding author on reasonable request.

### Study design

We conducted a cross-sectional study to examine whether *APOL1* genotype modified the relationship between smoking history and stroke prevalence in African American adults. At the time when participants were recruited, blood samples were collected and participants completed a questionnaire to document demographic characteristics, medical history (including history of stroke), and clinically important lifestyle factors.

### Ascertainment of Stroke

Briefly, the history of stroke was determined from the medical history reported in questionnaires. When available, medical records with neuroimaging (CT and/or MRI) and medical notes were retrieved and reviewed by a physician to further ascertain stroke type. Reported diagnosis with physician adjudication was used to classify the stroke events as an ischemic stroke or hemorrhagic stroke.

### Blood collection, lipid and lipoprotein analyses, and DNA preparation

Blood was collected after overnight fasting in tubes containing 0.1% ethylenediaminetetraacetic acid. When these samples were collected, blood was centrifuged at 3,000 rpm for 15 min at 4°C and plasma separated. An automated chemical analyzer (COBAS Chemistry analyzer) was used to measure levels of total cholesterol (TC), HDL cholesterol (HDL-C), and triglyceride (TG) in plasma as described previously.^21^ Very-low-density lipoprotein (VLDL) was prepared by ultracentrifugation.^22^ HDL-C was measured after precipitation of apoB-containing lipoproteins with dextran sulfate and magnesium.^23^ LDL cholesterol (LDL-C) was calculated as TC minus HDL-C plus VLDL-C or using the Friedewald equation when TG was <400 mg/dl. Genomic DNA was extracted using the Wizard purification kit (Qiagen).^*24*^ Both, DNA and plasma aliquots were stored at -80°C.

### Clinical and lifestyle covariates

Hypertension was defined as blood pressure ≥140/90 mm Hg or use of blood pressure– lowering medication. Type 2 diabetes mellitus (diabetes) was defined as fasting glucose ≥126 mg/dL or hemoglobin A1c ≥ 6.5% or use of diabetic mediation within 2 weeks prior to the clinic visit. Dyslipidemia was defined as age- and sex-normalized TG or LDL-C over the 90^th^ percentile, or HDL-C below 5^th^ percentile, or use of lipid–lowering medication. An exercise was defined as being active more than 30 min (including walking) more than twice per week, and alcohol consumption as having more than two drinks per week. Kidney disease is an independent risk factor for stroke. Carriers of two *APOL1* risk alleles have excess risk for kidney disease. To assess markers of kidney function, we measured plasma creatinine (mg/dL) following the manufacturer’s instructions (QuantiChrom™ Creatinine Assay Kit, BioAssay Systems, Hayward, CA).

### Smoking status

Smoking status was obtained from the self-reported smoking information in the questionnaire. Participants who answered yes to “Do you CURRENTLY smoke cigarettes?” were classified as *current* smokers. Participants who answered yes to, “Are you a PAST smoker?”, were classified as *past* smokers. Participants who responded no to these two questions were classified as *never* smokers. Cotinine, a main metabolite of nicotine, is the most commonly used biomarker of tobacco smoke exposure due to its relatively long half-life of ∼16 hours.^25^ To validate self-reported smoking status, plasma nicotine and cotinine levels were measured by gas chromatography (GS) at the UCSF Tobacco Biomarkers Core Facility. Briefly, concentrations of nicotine and cotinine were determined by gas chromatography with nitrogen-phosphorus detection,^26^ using 5-methylnicotine and 1-methyl-5-(2-pyridyl)-pyrrolidin-2-one (“ortho-cotinine”) as internal standards. This method has been modified for simultaneous extraction of nicotine and cotinine with determination using capillary GC.^27^ The limits of quantitation are 1 ng/ml for nicotine and 10 ng/ml for cotinine. The validation of smoking classification by questionnaires was assessed by comparison of self-reported smoking status with plasma nicotine and cotinine levels. From 485 available plasma samples, 60 (12%) participants had cotinine and/or nicotine levels detected. From 60 participants that had tobacco exposure markers in the plasma, 41 (68%) were self-reported *current* smokers, 13 (22%) self-reported *past* smokers, and 6 (10%) self-reported *never* smokers. All participants with detected cotinine and/or nicotine in plasma were considered *current* smokers.

### *APOL1* genotype status

PCR products were generated from primers [AAAACTGGCACGATAAAGGC and CATATCTCTCCTGGTGGCTG] designed using MacVector software (MacVector, Inc.).^24^ Sanger sequencing of exon 6 of *APOL1* gene was performed by the Quintara Biosciences. Sequencing chromatograms were analyzed using MacVector software (MacVector, Inc.) and *APOL1* genotypes determined separately by two persons. Participants’ *APOL1* genotypes were scored as G0/G0, G1/G0, G2/G0, G1/G1, G2/G2, or G1/G2. Based on scoring, participants were divided in two groups, *APOL1* risk genotype group and *APOL1* reference group. Participants were assigned to *APOL1* risk genotype group if having 2 risk alleles (G1/G1, G2/G2, G1/G2) or 1 risk allele (G1/G0, G2/G0, G0/G0), and to *APOL1* reference group if having 0 risk variants (G0/G0).

### Statistical Analysis

Statistical analyses were conducted using the RStudio (version1.3.1093) software. A significance level of α<0.05 was used to determined statistical significance. All variables were checked for missing values and skewness. Participants’ characteristics including demographics, medical history, smoking status and *APOL1* genotype status were examined and summarized for all participants, with respect to subgroups *ever* smokers vs. *never* smokers, and with respect to subgroups *APOL1* reference and *APOL1* risk genotype. The *P*-values and the descriptive statistics including the mean ± standard deviation (SD), median [range] for continuous variables or N (%) frequency distributions and percentages for binary and categorical variables are presented. A comparison of the distributions was performed using either an unpaired *t-*test for each continuous variable if variables passed normality distribution Shapiro tests, or using Wilcoxon rank sum tests if they were not normally distributed. Chi-squared test (Fisher’s exact tests) was used for each categorical variable.

We used logistic regression models to assess the relationship between smoking (*current* and *past* smokers vs. *never* smokers) and odds of stroke. All multivariable models were adjusted for important covariates. To control for potential confounding when examining smoking in relation to stroke, we included age, sex, dyslipidemia, hypertension, and diabetes in the final model. Body mass index (BMI), alcohol intake, exercise and plasma creatinine levels were also considered when building multivariable models. However, these covariates were not included in the final models because they did not meaningfully change the magnitude of the association between smoking and history of stroke, defined as a <10% change in the coefficient.

To test for effect modification by the *APOL1* risk genotype, we created an interaction term between the dichotomous smoking variable and the *APOL1* genotype variable (yes/no) and included the interaction term in the unstratified multivariate model. Given that the interaction term was significant, we reported the association between smoking and stroke stratified by the *APOL1* groups (*APOL1* reference group vs. *APOL1* risk genotype). Secondarily, we examined the dose-dependent effect of the *APOL1* genotypes on tobacco-related stroke. To do so, we examined the relationship between smoking and stroke among carriers of the *APOL1* reference genotype, carriers of one *APOL1* risk variant, and carriers of two *APOL1* risk variants separately. Thirdly, to evaluate the dose-dependent effect of smoking on history of stroke in *APOL1* groups (*APOL1* reference group vs. *APOL1* risk genotype), we assessed the association between smoking (*current* vs. *past*) and the odds of stroke for each of two *APOL1* groups individually. Lastly, in a sensitivity analysis, we assessed the association between smoking history and odds of ischemic stroke on a subgroup of 493 participants (we excluded 20 participants with no known stroke type).

## RESULTS

### Participants characteristics

Participants characteristics are shown in Table 1. Among all participants, after validating current smoking status by measuring tobacco exposure markers, 210 (41%) were *ever* smokers, with 70 (14%) *current* smokers and 140 (27%) *past* smokers. The average age of participants was 57.3 ± 13.9 years (range: 18 to 88 years). The *never* smoker group was younger, on average, than the *ever* smoker group (56.1 ± 14.8 years vs. 59.0 ± 12.4 years) with the majority being women in *never* smoker and men in *ever* smoker groups (57% and 45%, respectively). The average body mass index (BMI) was 28.6 ± 7.0 kg/m^2^ with no significant difference in BMI between *never* and *ever* smoker groups. The e*ver* smoker group had greater prevalence of hypertension, diabetes, and dyslipidemia (68%, 33%, 61%, respectively) than the *never* smoker group (46%, 23%, 41%, respectively). In contrast, the *never* smoker group had more physically active participants and fewer participants who reported drinking alcohol more than twice per week (63% and 12%, respectively) compared to the *ever* smoker group (43% and 20%, respectively). Among all participants, 54% had a *APOL1* risk genotype (1 or 2 risk alleles) with 12% having two *APOL1* risk alleles and 41% having one *APOL1* risk allele (Figure 1). Out of 41 stroke cases, 21 had available medical records, and stroke types were assessed. In all 21 stroke cases with available medical records, stroke types were determined to be an ischemic, and not hemorrhagic, strokes.

**Table 1.**
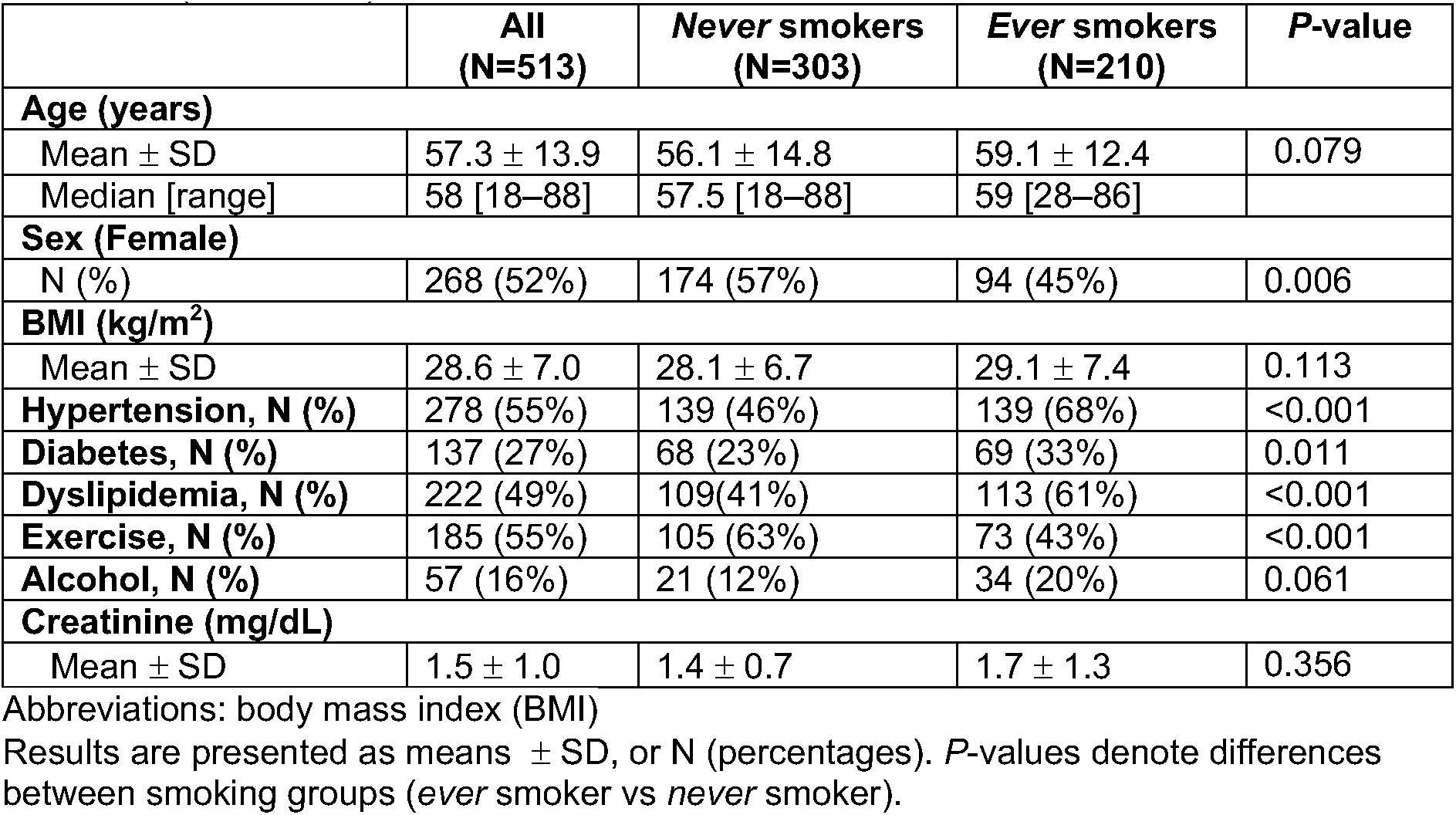
Characteristics of 513 African American adults in the cross-sectional study at enrollment (1999–2019)

**Figure 1.**
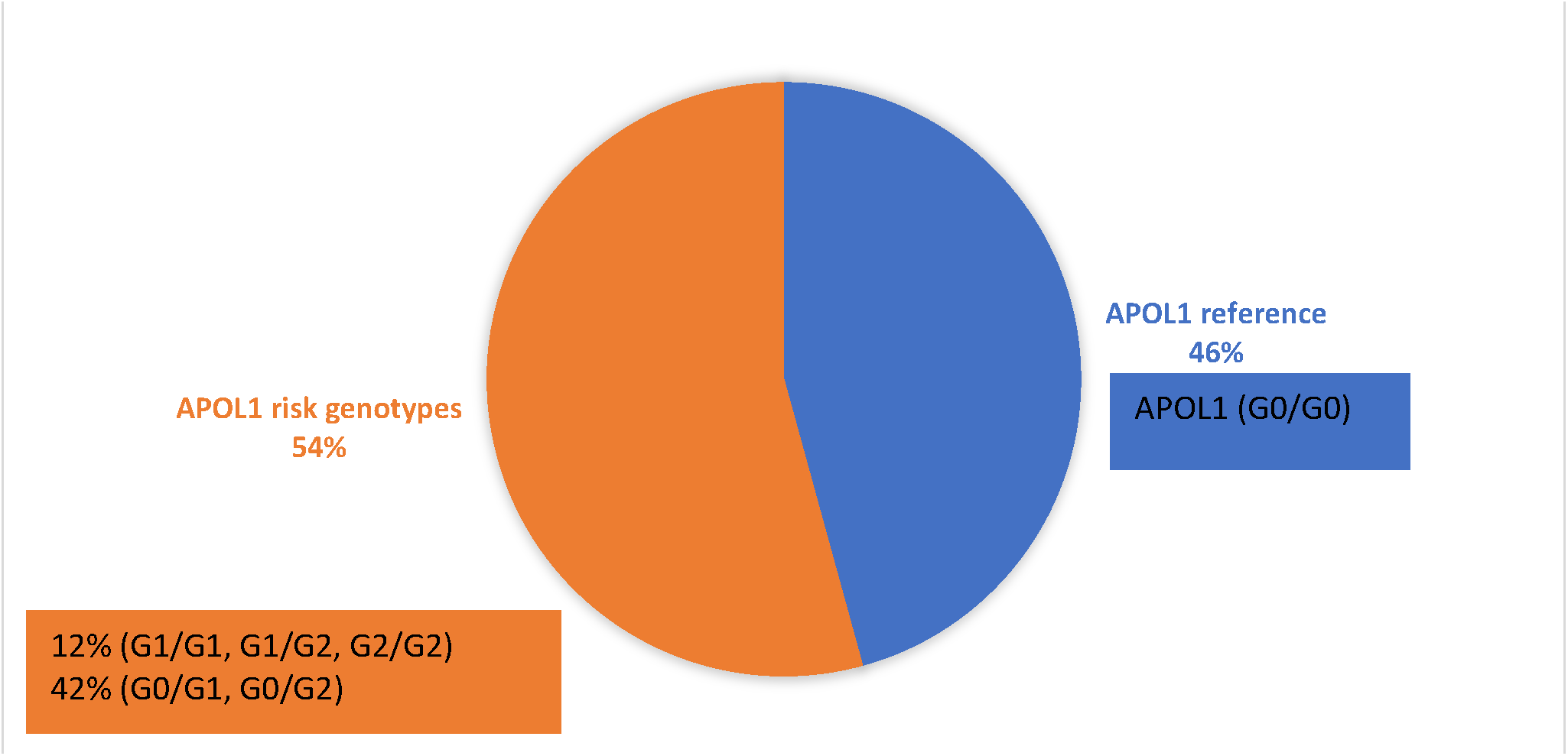
*APOL1* genotype frequency. Abbreviations: *APOL1* (Apolipoprotein L1)

### Prevalence of stroke in *never* and *ever* smokers differs by *APOL1* genotype

Overall, we identified 41 stroke events among all participants, 22 events (11%) among *ever* smokers and 19 (6%) among *never* smokers. Carriers of the *APOL1* reference genotype had similar prevalence of stroke events among *ever* and *never* smokers; 4 (4%) and 10 (7%) stroke events, respectively. Among carriers of the *APOL1* reference genotype, *ever* smokers had significantly greater prevalence of stroke compared to *never* smokers; 18 (15%) vs. 9 (6%) stroke events (Table 2). In a sensitivity analysis where participants with only ischemic stroke and no history of stroke were examined, we observed the similar findings where *ever* smokers had significantly greater prevalence of stroke compared to *never* smokers among carriers of the *APOL1* reference genotype, but not among non-carriers (Table 3).

**Table 2.**
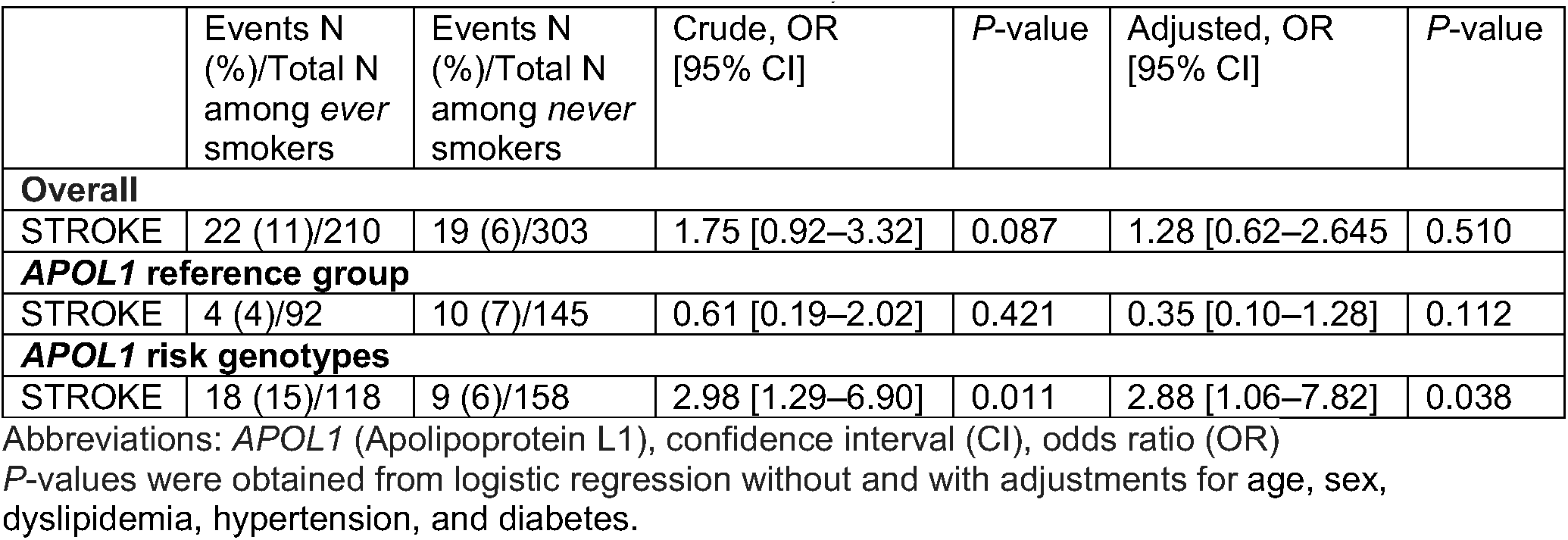
Prevalence, odds ratio [95% confidence interval] for stroke in *ever* smokers compared to *never* smokers among 513 African American adults, stratified by *APOL1* genotype status.

|  | Events N (%) / Total N among *ever* smokers | Events N (%) / Total N among *never* smokers | Crude, OR [95% CI] | P-value | Adjusted, OR [95% CI] | P-value |
| --- | --- | --- | --- | --- | --- | --- |
| **Overall** |  |  |  |  |  |  |
| STROKE | 22 (11)/210 | 19 (6)/303 | 1.75 [0.92–3.32] | 0.087 | 1.28 [0.62–2.645] | 0.510 |
| ***APOL1* reference group** |  |  |  |  |  |  |
| STROKE | 4 (4)/92 | 10 (7)/145 | 0.61 [0.19–2.02] | 0.421 | 0.35 [0.10–1.28] | 0.112 |
| ***APOL1* risk genotypes** |  |  |  |  |  |  |
| STROKE | 18 (15)/118 | 9 (6)/158 | 2.98 [1.29–6.90] | 0.011 | 2.88 [1.06–7.82] | 0.038 |

Abbreviations: *APOL1* (Apolipoprotein L1), confidence interval (CI), odds ratio (OR)
P-values were obtained from logistic regression without and with adjustments for age, sex, dyslipidemia, hypertension, and diabetes.

**Table 3.** Prevalence, odds ratio [95% confidence interval] for ischemic stroke in *ever* smokers compared to *never* smokers among 493 African American adults, stratified by *APOL1* genotype status.

|  | Events N (%) / Total N among *ever* smokers | Events N (%) / Total N among *never* smokers | Crude, OR [95% CI] | P-value | Adjusted, OR [95% CI] | P-value |
| --- | --- | --- | --- | --- | --- | --- |
| **Overall** |  |  |  |  |  |  |
| STROKE | 14 (7)/210 | 7 (2)/303 | 3.02 [1.20–7.63] | 0.019 | 1.62 [0.58–4.54] | 0.359 |
| ***APOL1* reference group** |  |  |  |  |  |  |
| STROKE | 1(1)/89 | 5 (4)/140 | 1.11 [0.29–4.27] | 0.874 | 0.47 [0.09–2.65] | 0.395 |
| ***APOL1* risk genotypes** |  |  |  |  |  |  |
| STROKE | 13 (12)/113 | 2 (1)/151 | 8.08 [1.73–37.65] | 0.008 | 5.00 [0.89–28.02] | 0.067 |

Abbreviations: *APOL1* (Apolipoprotein L1), confidence interval (CI), odds ratio (OR)
P-values were obtained from logistic regression without and with adjustments for age, sex, dyslipidemia, hypertension, and diabetes.

### Association of *APOL1* risk genotype with prevalence of tobacco-related stroke

In our crude and adjusted multivariate models including the interaction term (*ever* smoker\**APOL1* genotype) the association between smoking history and history of stroke differed by *APOL1* genotype in models (*P*^interaction^= 0.033, *P*^interaction^= 0.016, respectively), so we reported the data stratified by *APOL1* genotype in Table 2. In carriers of a *APOL1* risk genotype, *ever* smokers had 2.88 times the odds for a stroke compared to *never* smokers (95% confidence interval [1.06–7.82], adjusted *P* = 0.038). Relationships between smoking history and ischemic stroke, overall and stratified by the *APOL1* genotype status, are presented in Table 3. Among carriers of a *APOL1* risk genotype, *ever* smokers had 5.00 times the odds for an ischemic stroke compared to *never* smokers (95% confidence interval [0.89–28.02], adjusted *P* = 0.067).

In an effort to determine whether presence of two *APOL1* risk alleles has stronger effect on the associations between smoking and stroke than one *APOL1* risk allele, we performed secondary subgroup analyses. The OR for stroke comparing *ever* vs. *never* smoking increased with each increase in *APOL1* risk alleles (0 vs 1 vs 2) (Table 4).

**Table 4.** Odds ratio [95% confidence interval] for stroke in *ever* smokers compared to *never* smokers according to *APOL1* risk allele status (crude and adjusted models)

|  | ***APOL1* risk alleles** |  |  |
| --- | --- | --- | --- |
|  | **0 (N=238)** | **1 (N=212)** | **2 (N=63)** |
| STROKE (crude, OR [95% CI]) | 0.77 [0.25–2.32] | 2.51 [0.99–6.35] | 5.28 [0.56–50.20] |
| STROKE (adjusted OR [95% CI]) | 0.48 [0.15–1.59] | 2.35 [0.76–7.30] | 4.96 [0.31–80.13] |

Abbreviations: *APOL1* (Apolipoprotein L1), confidence interval (CI), odds ratio (OR)
Models (logistic regression) were adjusted for age, sex, dyslipidemia, hypertension, and diabetes.

### *APOL1* risk genotype in relation to stroke among those with different smoking history

*Current* smokers had 1.45 times greater odds that of the *past* smokers to report a history of stroke (95% confidence interval [0.47–4.48]). Among *APOL1* risk genotype group, *current* smokers had 1.20 times greater odds that of the *past* smokers to have stroke (95% confidence interval [0.30–4.76]). In the *APOL1* reference group, *current* smoker had 1.17 times odds that of the *past* smoker to have stroke (95% confidence interval [0.10–13.62]). All the results were non-significant (data not shown).

## DISCUSSION

In this study, the presence of the *APOL1* risk genotype modified the association between the smoking history and history of stroke in African American adults. Overall, *ever* smokers had numerically higher prevalence of stroke relative to *never* smokers. When the relationship between smoking and stroke was assessed in the *APOL1* subgroups separately, there was no association among non-carriers of the *APOL1* risk genotype. However, in carriers of *APOL1* risk genotypes, history of smoking was strongly and independently associated with greater odds of stroke. In particular, carriers of *APOL1* risk genotypes with a smoking history had almost 3 times the odds of having had a stroke compared to *never* smokers, after adjusting for covariates.

Furthermore, *ever* smokers who were carriers of *APOL1* two risk alleles tended to have greater odds for stroke than those who are carriers of *APOL1* one risk allele, though our cohort was not sufficiently powered to achieve statistical significance. Additionally, our sensitivity analysis implied the findings to be particularly true for ischemic type of stroke. Overall, our results suggest that carriers of *APOL1* risk genotypes may be susceptible to tobacco-related stroke, especially to ischemic stroke. Given high frequency of the *APOL1* risk genotypes in African Americans, this study may offer an explanation, at least partial, to excess burden of tobacco-related stroke recently reported in this high-risk population.

The presence of *APOL1* risk variants explains much of the excess risk for kidney disease observed in African American population.^9,12^ While some studies reported association between the *APOL1* risk variants and increased risk for stroke,^10,11,13^ other studies failed to confirm the association.^14-16^ The variability in results could be attributed to selection of study population and not being able to reach the potentially critical threshold for a development of stroke. Indeed, though exact mechanism is yet to be elucidated, findings from the animal study demonstrated that in addition to presence of *APOL1* risk genotype, the apoL1 risk variant expression levels are being causal in kidney disease with a particular emphasis on a “second hit” needed for rising apoL1 protein levels above a critical threshold leading to the development of the *APOL1* risk genotype related-kidney pathology.^17^ The importance of the “second hit” by an environmental trigger in *APOL1* risk variant-related kidney diseases was confirmed in the observational study.^28^ The similar mechanism could be relevant to *APOL1* risk variants role in tobacco-related stroke development. In our study we found notable interaction between *APOL1* risk variant and smoking history for stroke outcome, where smokers had higher odds for stroke only when they were carriers of the *APOL1* risk genotype.

Necrotic plaque formation is a result of inefficient removal of macrophages from arteries promoting the accumulation of cellular remains and extracellular lipids. Susceptibility to necrotic plaque formation and its enlargement due to apoL1 accumulation might be a biologic behavior among carriers of the *APOL1* risk variants. This notion is consistent with the findings from a transgenic mice study, where *APOL1* risk variants were found to promote cholesterol accumulation in tissues and macrophages due to downregulation of the major transporters involved in reverse cholesterol transport resulting in its impairment.^29^ Moreover, in a recent autopsy study, carriers of *APOL1 risk* genotypes compared to non-carriers had significantly larger necrotic cores of the plaques, and greater accumulation of apoL1 protein in the necrotic core in a gene-dose–dependent manner.^19^ Tobacco smoking is characterized by an increase in inflammation, with some inflammatory markers persisting more than five years after smoking cessation.^30^ Inflammation increases levels of apoL1 protein.^18^ Given that smoking-related inflammation may increase levels of apoL1 protein in the plaque’s necrotic core of the carriers leading to plaque disruption, it may be the “second hit” needed for stroke event occurrence, in particular ischemic stroke, explaining our observed associations. Indeed, we found that all stroke cases, for which medical records were available, were ischemic strokes. Additionally, the magnitude of the association became stronger in a crude and fully adjusted model with an ischemic stroke as an outcome. Overall, the relationship between smoking history and *APOL1* risk genotype demonstrated in our study underscores the notion that chronic inflammation initiated by tobacco smoking might be a trigger that leads to stroke, especially ischemic stroke, in susceptible individuals with *APOL1* risk genotype.

Tobacco smoking is a well-established risk factor for all forms of stroke.^3^ Smoking cessation reduces risk for stroke.^31^ While our sample size was limited, relative to *past* smokers, *current* smokers had higher prevalence of stroke, and this was independent of their *APOL1* genotype status. Risk for cardiovascular disease, including stroke, significantly declines within 5 years of smoking cessation.^31^ Although the *current* smokers were well-defined by plasma tobacco exposure markers, we did not have information on how many years had passed since smoking termination for our *past* smokers. Additionally, a recent study revealed a slow stroke risk decline beyond five years.^31^ Future larger studies with more comprehensive information on past smoking are needed to explore the role of the *APOL1* risk genotype in stroke risk among those with different history of smoking.

Our study has several limitations. First, our study was cross-sectional observational study and therefore our findings can only show an association, as opposed to direct proof of causal relationship. In addition, we were only able to examine odds of stroke events, rather than risk of incident stroke events. However, based on the biology described earlier, it seems plausible that the *APOL1* risk variants would also increase risk of incident stroke among *ever* smokers.

Second, as mentioned above, our exposure variable combines *current* and *past* smokers in one group. Future studies are needed to evaluate the role of *APOL1* genotype in stroke development among *current, past*, and *never* smokers separately, as well as among *past* smokers who quit smoking more than and less than five years ago. Third, our study population includes only self-reported African Americans, which limits our ability to generalize our findings beyond this population group. However, about half of African Americans are carriers of the *APOL1* risk genotypes, while it is very rare in other races indicating importance of understanding the role it plays in this high-risk population.

In conclusion, to the best to our knowledge, our study is the first study that has investigated the relationship between smoking and stroke in a self-reported African American cohort subdivided by *APOL1* risk genotype. We showed that the presence of the *APOL1* risk genotype modified the relationship between smoking history and stroke prevalence, particularly ischemic stroke, suggesting carriers of this genotype may be more susceptible to tobacco-related stroke. If confirmed in prospective cohorts, these findings could explain a large fraction of the uniquely high risk for stroke recently reported in African American smokers,^4^ and provide insights into critical targets for treatment that could help reduce racial stroke disparities.

## Data Availability

The data that support the findings of this study are available from the corresponding author on reasonable request.

